# Evaluating the impact of international airline suspensions on the early global spread of COVID-19

**DOI:** 10.1101/2020.02.20.20025882

**Authors:** Aniruddha Adiga, Srinivasan Venkatramanan, James Schlitt, Akhil Peddireddy, Allan Dickerman, Andrei Bura, Andrew Warren, Brian D Klahn, Chunhong Mao, Dawen Xie, Dustin Machi, Erin Raymond, Fanchao Meng, Golda Barrow, Henning Mortveit, Jiangzhuo Chen, Jim Walke, Joshua Goldstein, Mandy L Wilson, Mark Orr, Przemyslaw Porebski, Pyrros A Telionis, Richard Beckman, Stefan Hoops, Stephen Eubank, Young Yun Baek, Bryan Lewis, Madhav Marathe, Chris Barrett

## Abstract

Global airline networks play a key role in the global importation of emerging infectious diseases. Detailed information on air traffic between international airports has been demonstrated to be useful in retrospectively validating and prospectively predicting case emergence in other countries. In this paper, we use a well-established metric known as effective distance on the global air traffic data from IATA to quantify risk of emergence for different countries as a consequence of direct importation from China, and compare it against arrival times for the first 24 countries. Using this model trained on official first reports from WHO, we estimate time of arrival (ToA) for all other countries. We then incorporate data on airline suspensions to recompute the effective distance and assess the effect of such cancellations in delaying the estimated arrival time for all other countries. Finally we use the infectious disease vulnerability indices to explain some of the estimated reporting delays.

## OVERVIEW

For the ongoing COVID-19 epidemic, 24 countries^1^ had officially reported cases by the first week of February 2019. All of the first reports in these countries had travel history to China (mostly to Wuhan city or Hubei province). In this work, we assess the global importation risk of novel coronavirus disease (COVID-19) for other countries based on travel to China. We employ the concept of *effective distance* [3] which has been retrospectively validated for the SARS and H1N1 cases, to estimate the time-of-arrival (ToA) for all countries. We apply this metric on global traffic flows out of China obtained from the International Air Travel Association (IATA) [5] for the month of February 2019. We provide a risk assessment for countries based on two factors: the connectivity of the country to China (determined by the effective distance) and its vulnerability to disease outbreaks (determined by Infectious Disease Vulnerability Index (IDVI)).

We discuss the computation of the effective distance and the network construction in the subsequent sections. We show that with the constructed network, a moderately high linear relationship (coefficient of determination, *R*^2^ = 0.78) holds for ToA of COVID-19 cases as reported by World Health Organization (WHO) (Figure 1). We then employ the linear estimator to compute estimated ToAs for countries across the globe. By plotting the estimated ToAs against IDVI, a measure of vulnerability, we observe that of the countries that have reported cases, most are developed economies with typically high IDVI (low vulnerability) (Figure 2). This is likely due to (a) developed economies having strong air traffic and connectivity especially to China, and (b) their ability to detect and report imported cases fairly quickly. Our results indicate multiple countries (many with low IDVI, hence highly vulnerable) might see (or already have seen) case emergence in the month of February.

**Figure 1.**
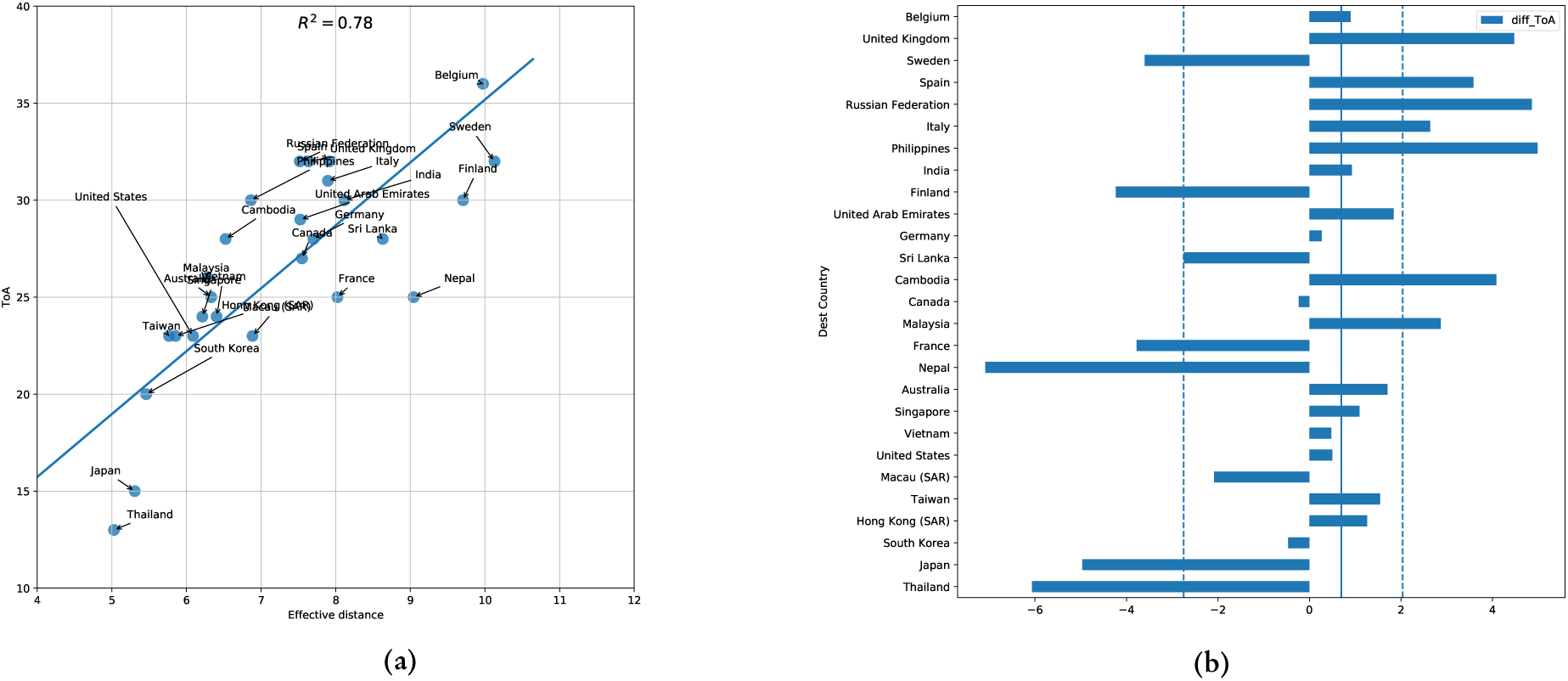
(a) Validating effective distance to the arrival times with first reports (Source: WHO Situation reports). Time of arrival is indexed starting Dec 31st, 2019 as day 0. (b) The difference between reported ToA and estimated ToA (with median error marked with solid vertical line and dashed lines indicating the interquartile range).

**Figure 2.**
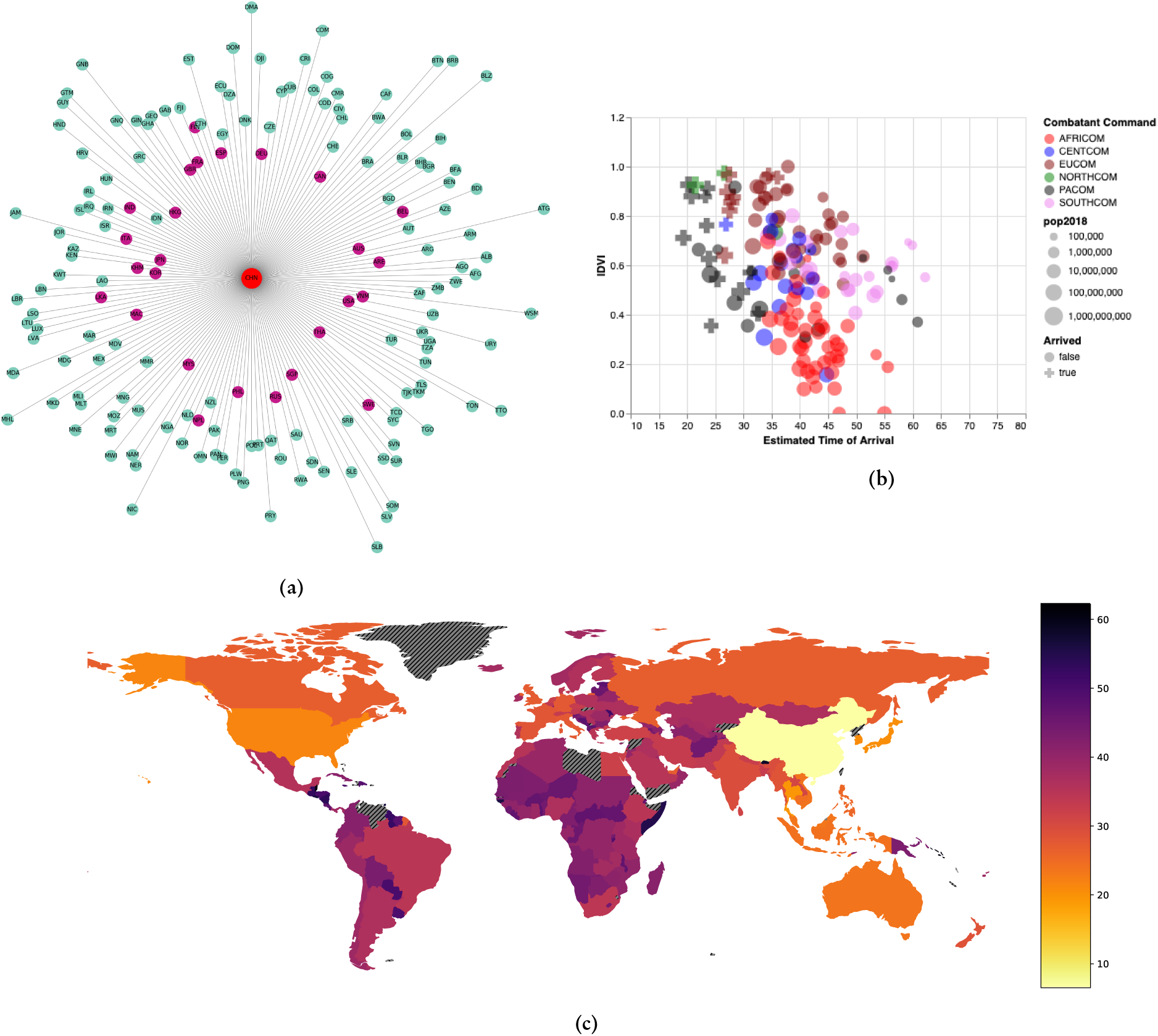
Effective distance and estimated arrival times to all countries. (a) A representation of the tree network with central node as China and the radial distance depicting the effective distance. (b) A country’s estimated ToA plottled against thier IDVI. Countries are colored by the US combatant command. Countries with official confirmed cases as of February 10th are shown with a plus marker. The full interactive visualization is available at https://nssac.github.io/covid-19/import_risk.html. (c) A choropleth plot of the countries on world map with colors indicating the estimated ToA. (slanted lines indicate countries with no direct airline links from China).

We then model the impact of flight suspensions to and from China that is being witnessed globally, while ensuring that the effective distances with and without interventions (i.e., suspensions) are comparable through normalization. The reduced flow from China results in a modified network with reduced edge weights and almost all countries see an increase in their effective distance from China (Figure 3), with some countries having higher increase than others. Finally, we also present an analysis comparing the difference in estimated versus reported time of arrival, with respect to the different component scores of IDVI (Figure 4), to investigate potential factors that may explain the reporting delays.

**Figure 3.**
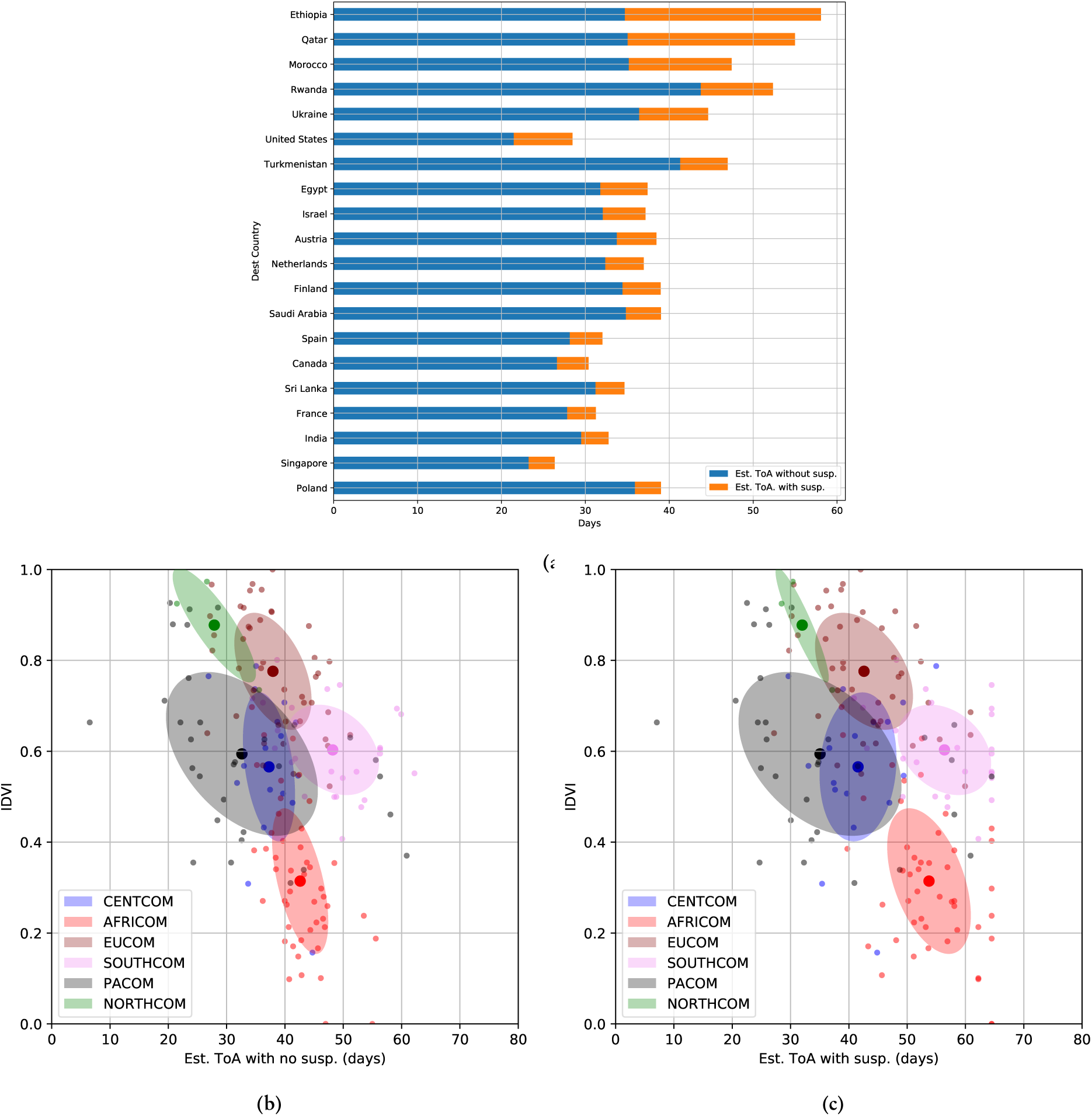
Change in estimated arrival times for various countries after incorporating airline suspensions (susp.) in the model. (a) Top 20 countries that see the highest change in est. ToA. IDVI for various countries plotted against their Est. ToA, (b) without suspensions (c) with suspensions. The mean-denoted ellipses indicate the standard deviation for the point clouds corresponding to the respective combatant commands.

**Figure 4.**
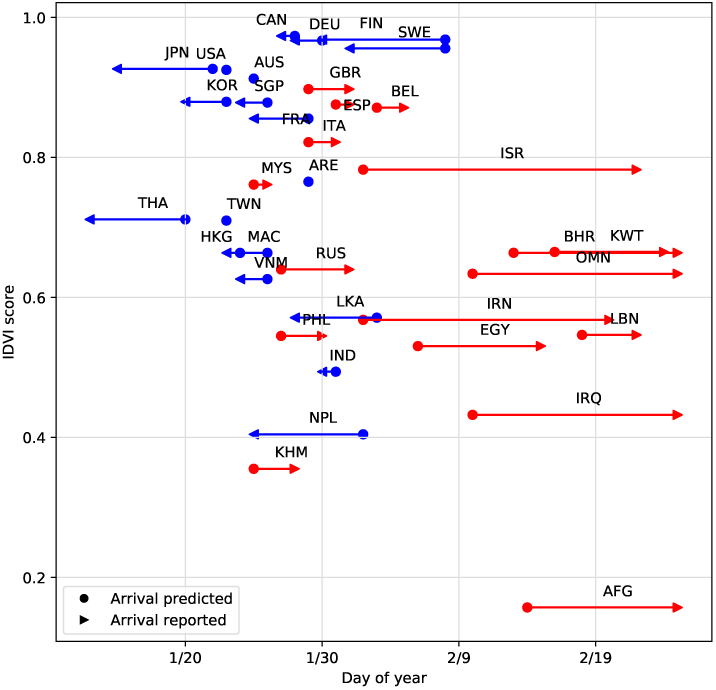
Comparison between estimated reporting delay and the overall normed IDVI score for countries reporting by February 25th 2020. Countries which reported later than estimated are colored in red, while those reporting earlier than estimated are colored in blue. The length of the lines represent the difference between reported and predicted ToAs.

### Related Work

There are multiple ongoing attempts to use airline traffic data to quantify global risk posed by COVID-19. In [1], the authors employ air travel volume obtained through IATA from ten major cities across China to rank various countries along with the IDVI to convey their vulnerability. [12] consider the task of forecasting international and domestic spread of COVID-19 and employ Official Airline Group (OAG) data for determining air traffic to various countries. [6] fit a generalized linear model for observed number of cases in various countries as a function of air traffic volume obtained from OAG data (authors refer to [12]) to determine countries with potential risk of under-detection. [7] provide Africa-specific case-study of vulnerability and preparedness using data from Civil Aviation Administration of China. In order to determine vulnerability of nations to COVID-19, the popular metric has been IDVI [1, 7] with [7] also employing WHO International Health Regulation Monitoring and Evaluation Framework to determine preparedness. For the current COVID-19 outbreak, [2] provides the *relative import risk* to various countries estimated using the world-wide air transportation network consisting of 4000 airports, and also an interactive visualization for arbitrary origin airports.

Some key differences: (a) We use IATA data which contains passenger flow volume between origin-destination airports (including transit points) instead of OAG data (used by [3]) which contains total number of seats available for a given segment. We believe that origin-destination flows by IATA provides a better estimate of population level exposure, and the likelihood of detecting the first case in a country (rather than at the transit airports), thus serves as a better network on which to compute effective distance (cf. Appendix A). While [1, 4, 8, 12] have used the IATA data for estimating relative risk for COVID-19 spread, this is the first work to show the relationship between the emergence times of COVID-19 using effective distance with actual arrival times for 24 countries. We provide estimated ToA for countries that have not officially reported COVID-19, and provide a way to quantify the effect of ongoing airline suspensions.

## METHODS

### Data

**IATA** [5] provides comprehensive air travel data covering more than 12000 airports across the world. Given a source and destination airport, it provides the number of passengers traveling between the two airports as reported by the airlines (“reported PAX”) and the IATA estimated (“reported + estimated PAX”) per month. The travel details include both direct and multi-hop connections between the airports. In addition, it provides the different carriers serving the two airports, which allow for simulating airline cancellations using historical data. For this analysis, we used data from the month of February 2019. **Infectious Disease Vulnerability Index** (IDVI) [9] is a metric developed by the RAND Corporation to identify countries potentially most vulnerable to poorly controlled infectious disease outbreaks because of confluence of factors including political, economic, public health, healthcare, demographics, and disease dynamics. **Arrival times** are curated from the official situation reports [10] published by World Health Organization. Each country is associated with the first date of confirmed case, with China being assigned December 31st, 2019. **Airline suspensions** information available as on February 10th 2020, by airline and routes was obtained and curated from Bloomberg [11] and other websites^2^.

### Network construction

We use international air traffic data from February 2019, which is the most recent data that coincides with the Lunar New Year period, known to involve a large movement of people in and out of China. As mentioned previously, IATA data provides number of passengers between two airports. We aggregate the flows out of all the airports in China to all the airports within the country of interest. We treat non-stop routes and routes with transit equivalently, since we are primarily interested in passenger volumes traveling out of China disembarking at various destination countries.

### Effective distance and estimated ToA

In order to model the risk due to direct importation, we consider the number of passengers whose trip originated in airport *m* and ended in airport *n* (through multiple paths) as obtained from IATA database. This data extracted from IATA is different from the number of available seats between *m* and *n* (i.e., the link capacity which could be used by passengers using *m → n* as a transit) considered in [3]. Hence, the computed effective distances would be different and the difference is explained in Appendix A. We compute the fraction of flows from China to any given country-of-interest and determine its effective distance. We develop a linear estimator for COVID-19 ToA at various countries using the effective distance to China. This allows us to “predict” when COVID-19 is most likely to be reported by various countries. Further, two countries with similar effective distances do not necessarily rank the same in terms of risk of sustained, undetected or uncontrolled epidemic outbreak. In order to capture this aspect, we consider the product of effective distance of a country to its IDVI to rank the countries by risk to COVID-19.

### Effect of airline suspensions

The rise in the number of COVID-19 cases in China has led to series of suspensed operations by major airlines serving international traffic in and out of mainland China. Note that airline suspension can be complete or partial, and can affect the whole or part of an itinerary. Using the suspension data described earlier, we appropriately alter the flow volumes in the original air traffic network. To ensure that the effective distances on the air traffic network are comparable with and without interventions, we scale the flow volume in the latter with total outflow from origin in the former. Assuming that the total outflow from a country without airline suspensions is reflective of the country population size, normalizing the reduced flows with it could provide better estimates for *P*_*mn*_. Formally, if *G* is the original weighted flow network on which effective distance is computed from source *i*_0_, and *G*′ is the flow network derived by adjusting flow volumes based on airline suspensions, then the edge weights between nodes *m* and *n* on *G*′ are 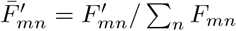, where 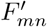 denotes the reduced flow between the nodes.

### Explaining estimated reporting delays

Finally, we seek to identify potential factors that can explain the *estimated reporting delays* according to our model. We define estimated reporting delay as the difference between the time of arrival predicted by the linear estimator and the reported time of arrival according to WHO situation reports. We construct univariate linear regression models with the IDVI scores (overall and individual categories), and report the % of explained variance according to each of these indicators. We also evaluated the goodness of fit via a Wald test with a t-distribution against a null hypothesis of a slope of 0 and report the p-values.

## RESULTS

### Importation risk prior to airline suspensions

Using a linear model for the reported time of arrival against the effective distance computed on IATA data, we observe a moderately high coefficient of determination *R*^2^ = 0.78. A scatter plot of COVID-19 ToA for countries with reported cases with respect to their effective distance from China is shown in Figure 1a. It should be noted that in the regression analysis, China has effective distance and time of arrival set to zero. The difference between reported ToA and linear estimator is provided in Figure 1b. The median error in estimation is less than a day with the estimated ToA typically being earlier than the reported ToA. Note that there are a few outliers which have much earlier reported ToA than estimated ToA (e.g., Nepal, Japan, Thailand). For these countries, airline traffic may not be representative of the total connectivity with China due to their spatial proximity (land or sea). In order to assess the risk of direct importation of COVID-19 from China to various countries we computed the estimated ToA for each country, using their effective distance from China (shown in Figure 2). The effective distance tree plot ^3^ clearly show that the early reporting countries indeed had the shortest effective distance to China (at the center). The predicted time of arrival using the linear estimator is shown in the map, excluding countries which do not have direct airline traffic from China. Finally, we plot the estimated times of arrival of all countries against their vulnerability (IDVI), grouped by the United States Combatant Commands, providing a natural geographic grouping. As demonstrated in Figure 2b, the countries with reported cases (shown with plus marker) are mostly to the left. Interestingly, most countries with reported cases also have higher IDVI, which may be due to a combination of (a) high connectivity and air traffic to China, (b) better detection and reporting capabilities for imported cases. In general, the lower left region (low IDVI and low estimated ToA) indicates a regime of high risk, which is relatively empty for this outbreak. However, we notice that several low IDVI countries (especially those in AFRICOM) are estimated to have times of arrival in the first three weeks of February. Equally concerning are countries to the left with circle markers, which have an early estimated time of arrival but haven’t officially reported cases yet.

### Impact of airline suspensions

For a select set of countries, we compute their effective distance with and without airline suspensions and subsequently, the estimated ToA for each case. The top 20 countries as per the change in estimated ToA after suspension are presented in Figure 3a. Since our network alteration ensures monotonicity after flow volume reduction, we observe that a decrease in flow volumes results in increased estimate for ToA across all countries. We observe that most countries (Figure 3a), on an average, see an increase in estimated arrival time of *≈* 4 *−* 5 days, while, Ethiopia and Qatar observe an increase *>* 10 days.

We select the countries for which there have been no imported cases, and compute their new estimated time of arrival using the updated effective distance. For this process, we use the regression coefficients that were computed using known arrival times. Figures 3b and 3c show the scatter plots of estimated ToA vs. IDVI for the countries before and after the imposition of airline suspensions. The ellipses indicate the *σ−*confidence interval of the point clouds corresponding to the respective combatant commands. The increase in ellipse widths and mean shift to the right (due to constant IDVI) between Figure 3b and Figure 3c indicate the effect of flight cancellations on estimated ToA. On an average, countries in AFRICOM see the largest average increase in estimated ToA of nearly 11 days, followed by countries in SOUTHCOM showing an average increase of 8 days.

## ESTIMATED REPORTING DELAY AND IDVI

We recall that the output of our linear estimator is the expected ToA based on airline flow. We computed the estimated reporting delay for countries reporting before February 26th 2020 (36 of them according to WHO situation reports). Figure 4 shows the reported and predicted ToA for these countries, colored based on whether they were earlier (blue) or later (red) than predicted. The results of univariate regressions of each of the IDVI scores is shown in Table 1. We generally observe the negative relationship between the reporting delays (lower IDVI leads to higher reporting delays). Of the different normed scores, we see that the political scores (domestic and international) have the highest coefficients of determination (24% and 18% respectively, *p <* 0.05). The domestic score includes as components indicators of governance, corruption, decentralization, government stability, etc. while the international score captures aspects such as collaboration, international organization support and aid dependence. These are aspects which could have direct or indirect bearing on the detection and reporting of infectious disease outbreaks. The overall IDVI score explains about 16% of the variance (*p <* 0.05). While this is a reasonable first step, and does explain some of the apparent late reporting we are observing, we note that the estimated reporting delay may need to be replaced by other metrics of reporting deficiency derived from case surveillance, as new countries continue to report cases of COVID-19.

**Table 1.** Relationship between IDVI scores and projected reporting delays. Overall IDVI explains about 16% of the variance in estimated reporting delays, while political domain scores (domestic and international) explain about 24% and 18% respectively (*p <* 0.05).

| Score | Slope | $R^2$ | p-value | std-error |
| --- | --- | --- | --- | --- |
| Overall IDVI | -14.70 | 0.16 | 0.02 | 5.78 |
| Demographic | -14.26 | 0.11 | 0.05 | 7.11 |
| Healthcare | -12.82 | 0.06 | 0.15 | 8.63 |
| Public health | -12.28 | 0.08 | 0.10 | 7.32 |
| Disease dynamics | 6.65 | 0.02 | 0.40 | 7.96 |
| Political (domestic) | -13.35 | 0.24 | 0.002 | 4.09 |
| Political (international) | -14.24 | 0.18 | 0.01 | 5.24 |
| Economic | -7.23 | 0.05 | 0.19 | 5.43 |

## DISCUSSION

Our work is an initial attempt at quantifying the impact of airline suspensions on COVID-19 direct importation risk. Some of the limitations of the current work can be overcome with more timely data availability and improved model assumptions. Firstly, the current observations are based on first official reports which in some cases can be quite different from first importations. Also, due to the evolving nature of the outbreak and limited observations, the linear estimator coefficients could change with new reports and altered travel conditions.

While air traffic data from IATA allows us to quantify population exposure, (a) it is dated and may not be reflective of current conditions; (b) may not be representative of all human mobility between the countries. As we observed, some countries that are geographically closer to China (e.g., Nepal, Thailand and Japan) have very early arrival times (in relation to estimates based on effective distance). This highlights the need to account for multi-modal transport networks for quantifying the risk of global importations. However, this also raises concerns about other countries and regions (such as Pakistan, Myanmar and Northeastern India) which are geographically adjacent to China but haven’t reported any cases yet. Notably, many of these regions scored relatively poorly on the indices of societal function, suggesting ties between the openness of a given regional government and the timeliness and accuracy of their data.

Finally, we have considered China as a single origin, while there exists case counts by province. The current analysis can be extended by considering weighting the effective distance from multiple origins with their relative levels of infection. Further, while we have tried to incorporate data on airline suspensions, this may not be complete, and will not capture the actual reductions in flow volumes due to travel advisories, government restrictions and behavioral adaptations. There are also a number of screening procedures in place at international airports, which could potentially delay case importations.

While providing a preliminary analysis using COVID-19 official reports and airline suspensions, this work also lays out a framework for rapid risk assessment for an emerging and ongoing outbreak. We believe that using near real-time multi-modal mobility datasets and detailed disease surveillance with qualitative and quantitative inputs on ongoing interventions and preparedness efforts will aid in swift and efficient global response to such outbreaks.

## Data Availability

All data except from proprietary sources used in the analysis are made available as Supplemental material.

https://doi.org/10.18130/V3/Z6524P

## ACKNOWLEDGMENTS

The authors would like to thank members of the Network Systems Science and Advanced Computing (NSSAC) Division for interesting discussion and suggestions related to epidemic science and machine learning. This work was partially supported by National Institutes of Health (NIH) Grant 1R01GM109718, NSF BIG DATA Grant IIS-1633028, NSF DIBBS Grant ACI-1443054, DTRA subcontract/ARA S-D00189-15-TO-01-UVA, NSF Grant No.: OAC-1916805, US Centers for Disease Control and Prevention 75D30119C05935, and a collaborative seed grant from the UVA Global Infectious Disease Institute.

## Appendix

### A EFFECTIVE DISTANCE COMPUTATION OVER IATA-BASED AND OAG-BASED NETWORKS

Consider the two networks presented in Figure 5. Here *m, 𝓁*_*a*_, *𝓁*_*b*_, *𝓁*_*c*_, and *n* represent the nodes and the effective distance is computed between node *m* and *n*. Figure 5a represents the network construction using IATA data for computing the effective distance between node *m* and *n*. In this construction, *f*_*mn*_ represents the number of passengers (passenger flow) on the path [(*m, n*)]. IATA data consists of all the paths (along with the flow) that were used by passengers in the travel between source *m* and destination *n*. In the example network (Figure 5a), the paths are Γ = *{*[(*m, n*)], [(*m, 𝓁*_*a*_), (*𝓁*_*a*_, *n*)], [(*m, 𝓁*_*b*_), (*𝓁*_*b*_, *𝓁*_*c*_), (*𝓁*_*c*_, *n*)]*}* with respective flows as 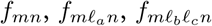 The total flow between nodes *m* and *n* is the sum of the flows through all the paths connecting them and we denote it as 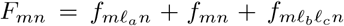 With ∑_*k*_ *F*_*mk*_ as the total outflow from node *m*, we define 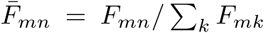, as the probability of a traveller exiting node *m* has destination node *n*. The effective distance between nodes *m* and *n* is defined as 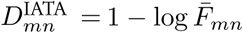.

**Figure 5.**
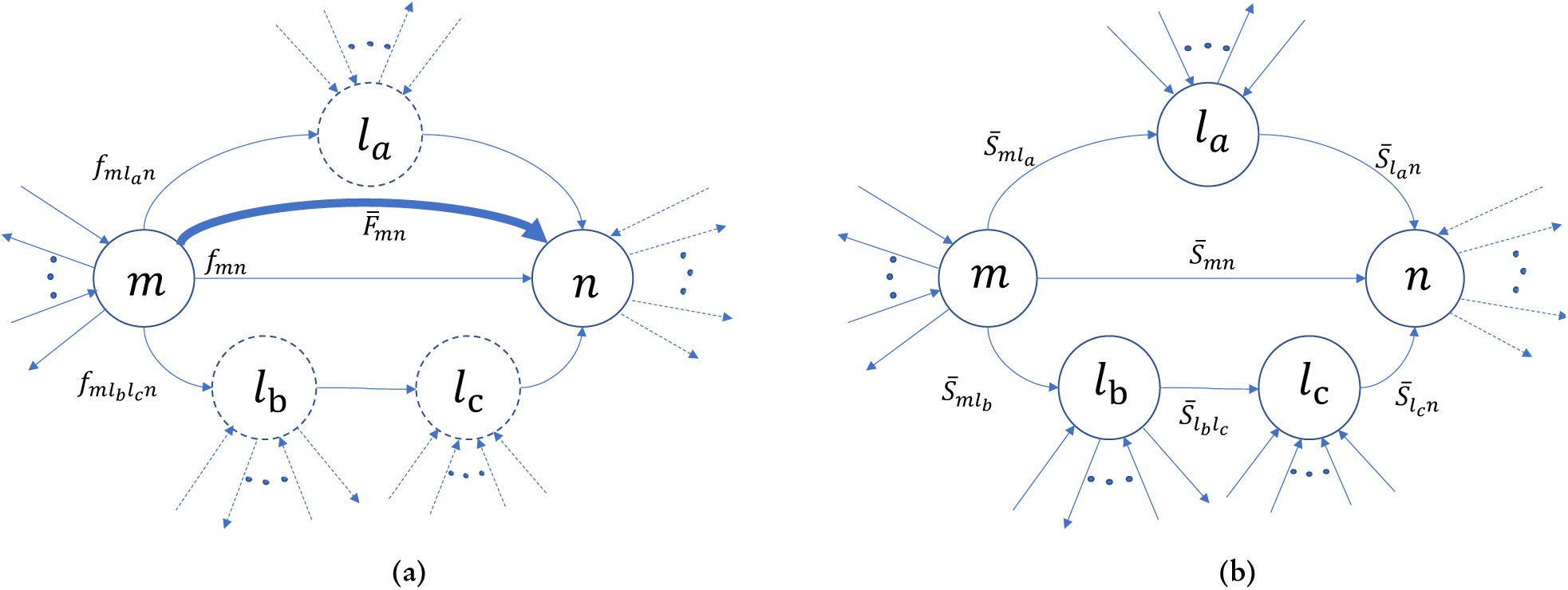
A toy example that compares networks derived from IATA and OAG data. Nodes *n* and *m* are considered as source and destination, respectively. (a) Network constructed using IATA data 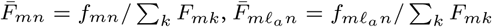, and 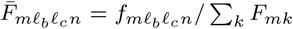, the bold, arrow is used to represents 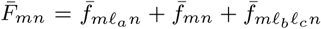, the total fraction of flow from origin *m* to destination *n*. (b) Network constructed using the OAG data. The dashed lines are used to represent nodes and outflows that do not contribute to the computation of the edge weights in the graph.

As for network construction using OAG data (cf. Figure 5b) we denote *S*_*mn*_ as the total number of seats between airports *m* and *n*. 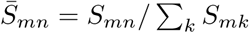 denotes the probability of a person exiting node *m* (as either origin or transit) has the next stop at node *n* and the effective distance is defined as 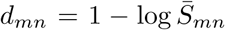. In [3], the authors consider all the paths that can be constructed between the two nodes which in this case is the set Γ and define the effective distance as the shortest path between the two nodes: 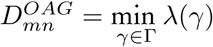, where *λ*(*γ*) = ∑_(*i,j*)*∈γ*_*d*_*ij*_ is the length of path *γ*.

## Footnotes

1 excluding Hong Kong, Macau, and Taiwan. 33 countries as of 25th February 2019 according to WHO situation reports

2 https://www.airchina.us/,https://www.hainanairlines.com/,https://www.oag.com/

3 Figure generated using https://github.com/benmaier/radial-distance-layout

